# Body composition predicts poor outcomes and reveals immunometabolic dysfunction via single-cell profiling in anti-BCMA CAR T-treated myeloma

**DOI:** 10.1101/2025.08.23.25334251

**Authors:** Thomas C. Wiemers, Michael Rade, Nora Grieb, Maximilian Ferle, Tihomir Dermendzhiev, David Fandrei, Patrick Born, Luise Fischer, Sabine Seiffert, Anja Grahnert, Maik Friedrich, Ronny Baber, Markus Kreuz, Klaus H. Metzeler, Marco Herling, Carmen D. Herling, Madlen Jentzsch, Georg-Nikolaus Franke, Andreas Boldt, Thomas Neumuth, Urvi A. Shah, Ulrike Köhl, Kristin Reiche, Timm Denecke, Uwe Platzbecker, Vladan Vucinic, Hans-Jonas Meyer, Maximilian Merz

## Abstract

Chimeric Antigen Receptor (CAR) T-cell therapy has transformed the treatment of relapsed or refractory multiple myeloma (RRMM), yet outcomes remain heterogenous. The prognostic role of body composition in this context is unknown. We retrospectively analyzed 108 RRMM patients treated with anti-B-cell maturation antigen (BCMA) CAR T-cell therapy. Pre-treatment Computed tomography imaging was utilized to quantify total adipose tissue (TAT), subcutaneous adipose tissue (SAT), visceral adipose tissue (VAT), and skeletal muscle area to assess sarcopenia. Longitudinal flow cytometric and single-cell multi-omic analyses were conducted to characterize the quantitative and qualitative influences of body composition on the immune microenvironment. Patients with BMI <25 kg/m^2^ experienced significantly worse overall survival (OS) compared to high-BMI patients. Reduced TAT, primarily driven by low SAT, was associated with inferior OS, diminished response and elevated soluble BCMA. Sarcopenia independently predicted poorer OS, while progression-free survival was unaffected by the respective parameters. Low SAT and sarcopenia correlated with lower bystander T-cell counts at leukapheresis. Longitudinal T-cell receptor sequencing and single-cell transcriptomics revealed diminished cytotoxic and interferon signaling, reduced T-cell clonality, and increased oxidative phosphorylation activity following CAR T-cell infusion. Our findings identify low SAT and sarcopenia as prognostic biomarkers that influence survival, therapeutic response, and immunometabolic profiles. Their quantification through standard imaging techniques offers a cost-effective strategy for early risk stratification and individualized management in CAR T-cell therapy.

## Introduction

Chimeric Antigen Receptor (CAR) T-cell therapy has revolutionized the treatment landscape for relapsed or refractory multiple myeloma (RRMM), offering durable responses even in heavily pretreated patients (1). Despite these transformative advances, outcomes vary due to factors like age, comorbidities, prior therapies, high-risk cytogenetics, and extramedullary disease (EMD) (2,3). Additionally, intrinsic features of the CAR T-cell product - such as T-cell phenotype, and in vivo expansion kinetics - play critical roles in shaping therapeutic efficacy (4,5). Recent studies have also highlighted the prognostic relevance of early treatment response, as well as the composition and dynamics of the non-transduced T-cell compartment and immunosuppressive bystander cells (6). While these clinical and cellular determinants are increasingly well characterized, the role of body composition particularly sarcopenia and adipose tissue distribution remains underexplored in the context of CAR T-cell therapy. Existing data from patients with CD19-directed CAR T-cell therapy suggest that sarcopenia may adversely impact outcomes, possibly through reduced physiologic reserve, heightened treatment-related toxicity, or impaired immune responsiveness (7,8). In solid tumors, improved outcomes following immunotherapy have been associated with greater skeletal muscle mass and regular exercise, pointing to a potential link between physical fitness, immune competence, and therapeutic response (9–12). In multiple myeloma (MM) the prognostic relevance of body composition remains uncertain (13–15). Emerging evidence suggests that obesity may increase the risk of monoclonal gammopathy of undetermined significance (MGUS) and influence disease progression, but its impact on treatment response is less clear (16). To date, no studies have systematically examined how fat distribution or sarcopenia affect outcomes after CAR T-cell therapy in MM, which represents a crucial gap as physical activity and nutrition interventions are explored (10,17).

To address this, we conducted a large real-world study examining the prognostic significance of computed tomography (CT)-derived body composition parameters including total adipose tissue (TAT), subcutaneous adipose tissue (SAT), visceral adipose tissue (VAT), and skeletal muscle area (SMA) in patients with RRMM undergoing anti-B-cell maturation antigen (BCMA) CAR T-cell therapy. Beyond baseline assessment, we integrated these findings on CAR T-cell expansion and longitudinal data on bystander immune cell dynamics, leveraging a high-resolution single-cell RNA and T-cell receptor (TCR) sequencing dataset to provide a comprehensive systems-level view of how body composition influences immune fitness and therapeutic outcomes after CAR T-cell therapy in MM.

## Methods

### Patients and assessments

We enrolled 108 patients with relapsed or refractory multiple myeloma (RRMM) who received the commercially available CAR T-cell therapies idecabtagene vicleucel (Ide-cel) or ciltacabtagene autoleucel (Cilta-cel) outside of clinical trials at the University Hospital Leipzig, Germany. The study adhered to the Declaration of Helsinki and was approved by the University Hospital Leipzig ethics committee (361/22-ek). All participants provided written informed consent and received no financial compensation. CAR T-cells were administered intravenously three to five days after lymphodepleting chemotherapy (LDC), in accordance with the manufacturer’s instructions. For risk assessment prior to CAR T-cell infusion, refractoriness, presence of EMD as well as revised International Staging System (R-ISS) and Endothelial Activation and Stress Index (EASIX) scores were assessed (18–20). The occurrence of cytokine release syndrome (CRS) and Immune Effector Cell-Associated Neurotoxicity Syndrome (ICANS) was documented and classified according to the consensus criteria established by the ASTCT (21). Response assessment was performed around 30- and 100-days post-infusion and categorized in accordance with IMWG criteria as progressive disease (PD), stable disease (SD), partial remission (PR), very good partial remission (VGPR) or complete response (CR) (22).

### Imaging studies and muscle/adipose tissue quantification

CT imaging was conducted using a clinically employed CT scanner with either 128 or 256 slices (Ingenuity or iCT256, Philips, Hamburg, Germany). For determination of all subsequent parameters, we used 2D CT slices from the region of L3. These slices were obtained from routinely acquired CT scans before CAR T-cell therapy that are part of the standard protocol for MM surveillance (22) (**Figure 1a**). The volume of the cross-sectional area was assessed in accordance with previous studies (13,23,24). SMAs of psoas muscle, paraspinal muscles and abdominal wall muscles were determined semiautomatically using ImageJ software 1.48 (National Institutes of Health Image program, USA) within a range of –29 and 150 HU. Skeletal muscle index (SMI) was calculated with SMA in relation to body weight. Similarly, VAT and SAT were calculated semiautomatically using a threshold of –190 and –30 HU, respectively. TAT represents the combined total of both fat areas.

**Fig. 1.**
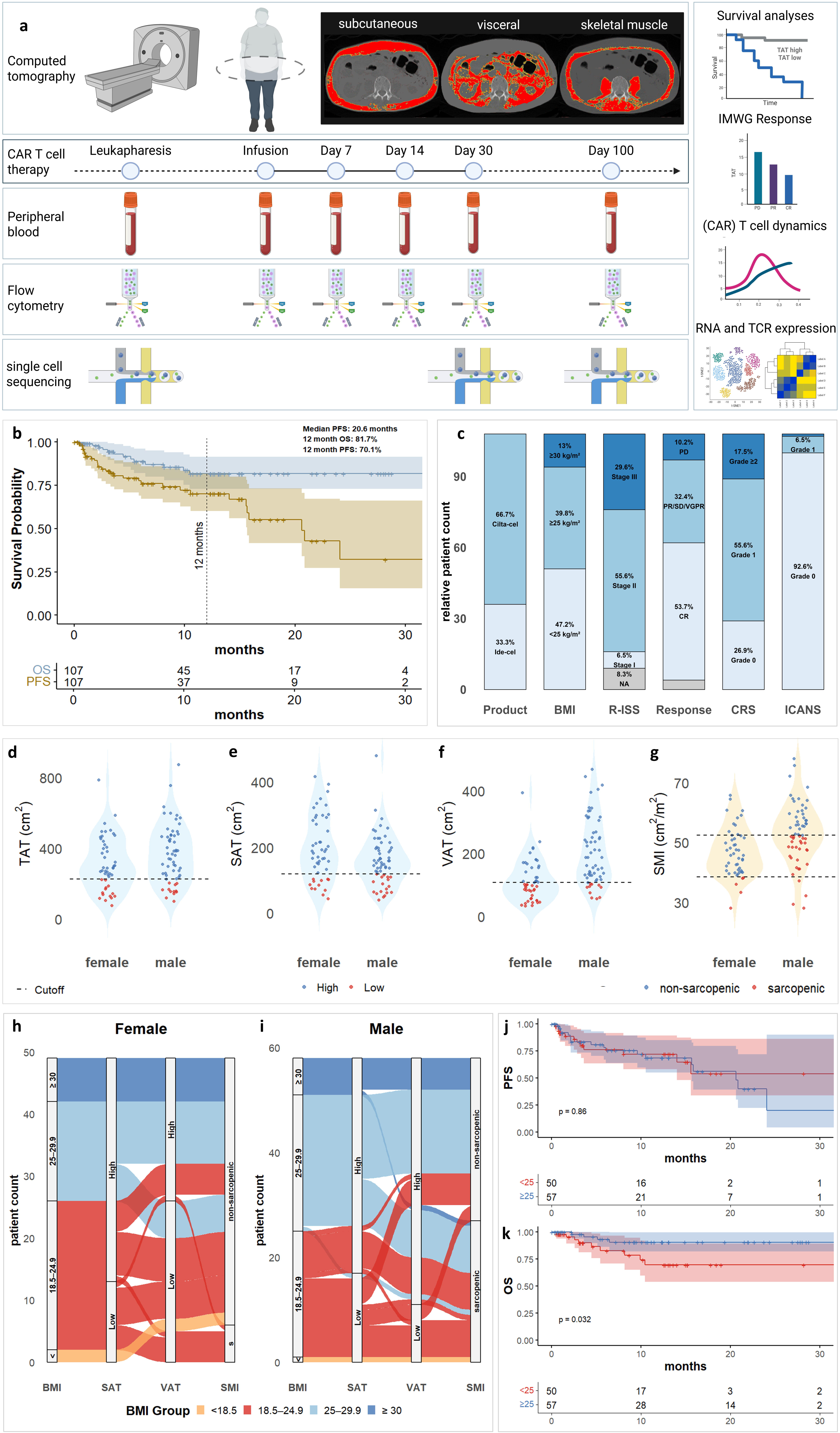
Study Workflow. **a**, Overview of the study design showing computed tomography (CT) imaging for body composition analysis, the CAR T-cell therapy timeline, and sample collection for peripheral blood, flow cytometry, and single-cell sequencing, with downstream analyses including survival curves, IMWG response, CAR T-cell dynamics, and RNA/T-cell receptor(TCR) profiling. **b**, Kaplan-Meier curves display overall survival (OS, blue) and progression-free survival (PFS, green) with 12-month OS and PFS rates and numbers at risk indicated. **c**, Clinical and treatment characteristics such as CAR T-product, BMI categories, R-ISS stage, response rates, CRS, and ICANS grades are summarized. **d-g**, Violin plots illustrate total adipose tissue (TAT), subcutaneous adipose tissue (SAT), visceral adipose tissue (VAT), and skeletal muscle index (SMI), stratified by sex and grouped by high/low or sarcopenic/non-sarcopenic status, with horizontal dashed lines marking cutoff values. **h-i**, Sankey diagrams show BMI categories, SAT, VAT, and SMI distributions in female and male patients. **j-k**, Kaplan-Meier curves for PFS and OS compare BMI groups (<25 vs. ≥25 kg/m²), with log-rank test used for statistical analysis. Shaded regions represent 95% confidence intervals, and numbers at risk are indicated below each plot.

### Sample collection and processing

To analyze the impact of CT-derived body composition on immune cell composition and soluble BCMA (sBCMA), peripheral blood samples were collected on the day of Leukapheresis (LP), the day of CAR T-cell infusion (day 0), and on days 7, 14, 30, and 100 post-infusion (**Figure 1a**). Flow cytometric analysis was performed promptly on fresh samples using antibody panels to identify CAR^+^ T-cells and to study the T-cell compartment, as described previously (5,25). Samples were processed with standardized staining, lysis, and washing steps, and analyzed using a BD FACSLyric™ system. In addition, we performed single-cell RNA and TCR sequencing on Peripheral Blood Mononuclear Cells (PBMCs) collected on the day of LP, as well as at Late (day 19–64, average of 31) and Very late (day 63–169, average of 102) time points following CAR T-cell infusion. For the analysis, we followed our previously established protocol (5).

### Statistical analysis

All analyses were conducted using R (v4.5.0). Cox proportional hazards models assessed associations between variables and survival outcomes. For survival analyses, including hazard ratios and log-rank tests, survminer (v0.0.5) and survival (v3.8.3) package were used. To determine optimal cut-off values for adipose tissue parameters, we stratified the cohort into two groups by maximizing the log-rank statistics for outcomes associated with mortality (maxstad-method), which were subsequently applied in all survival outcome analyses. These values closely align with thresholds reported by other groups employing different methodological approaches (13,14). Sarcopenia was defined based on established cut-offs by Prado et al. (23). Kaplan-Meier survival analyses were performed starting from the time of CAR T-cell therapy to the progression of MM (progression-free survival, PFS) or to death from any cause (overall survival, OS). Differences in cell type proportions and patterns of differential gene expression were analyzed as described previously (5). Statistical significance was set at p < 0.05, unless otherwise specified, using log-rank, Chi-Square and Wilcoxon rank-sum tests when appropriate. For longitudinal group comparisons, p-values were adjusted for multiple testing at each time point using the Benjamini–Hochberg method.

## Results

### Obesity and Sarcopenia are common in RRMM treated with CAR T-Cell Therapy

The cohort consisted of 108 patients with a median age of 64.3 years at CAR T-cell infusion, including 72 (66.7%) treated with Cilta-cel and 36 (33.3%) with Ide-cel. For one patient (0.9%), no CT scan was available prior to treatment. A workflow that introduces the principal steps and sample collection within our study is presented in **Figure 1a**. Median PFS was 20.6 months, while median OS was not reached, with 12-month PFS and OS rates of 70.1% and 81.7%, respectively (**Figure 1b**). CRS occurred in 79 patients (73.1%), predominantly grade 1 in 60 (55.6%) patients, and grade 2 or higher in 19 patients (17.5%) (**Figure 1c**).

The median body mass index (BMI) was 25.3 kg/m², with 3 (2.7%) patients having a BMI < 18.5 kg/m², 47 (43.5%) between 18.5 and 24.9 kg/m², 44 (40.7%) patients between 25.0 and 29.9 kg/m², and 14 (12.9%) patients ≥ 30.0 kg/m². The median age at CAR T-cell infusion was 64.3 (IQR 10.6) years. Median TAT, SAT, and VAT were 309.2, 161.0, and 137.6 cm², respectively, and the median SMA was 142.9 cm². In the entire cohort, 27 (25%), 30 (27.8%), and 37 (34.2%) patients were identified as low TAT, SAT, and VAT, respectively, and 33 patients (30.5%) were classified as sarcopenic (**Figure 1d–g**). A complete response was achieved in 58 patients (53.6%). Body composition parameters were independent of EASIX, R-ISS, CAR T-cell product, age prior to infusion and occurence of CRS. Non-relapse mortality (NRM) was observed in 5 (4.6%) patients without correlation to adipose tissue parameters or sarcopenia. Notably, low SAT was associated with elevated sBCMA levels at LP, day 0, and day 30 post-infusion. Detailed baseline characteristics are presented in **Table 1**.

**Table 1:**
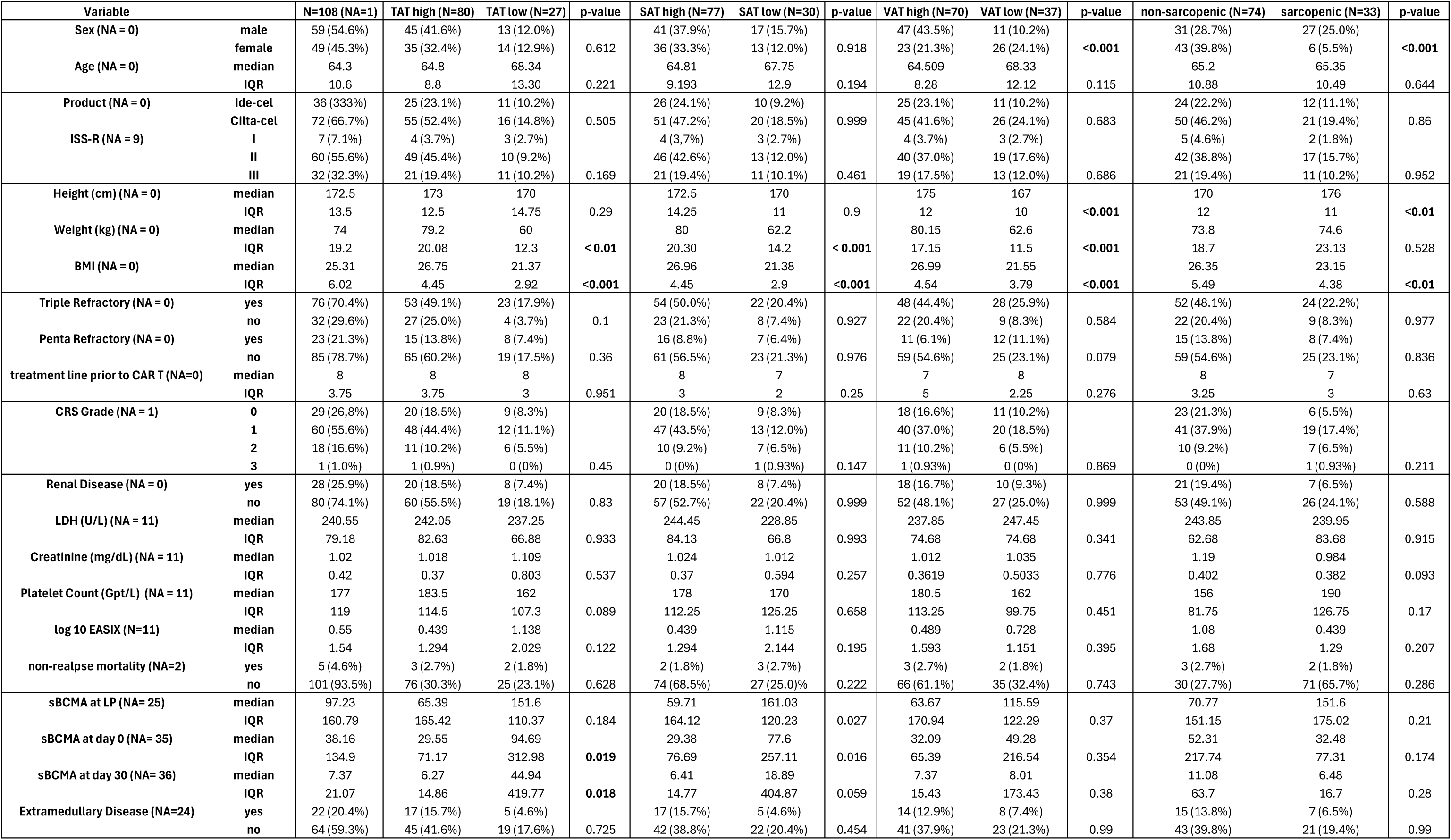
Baseline Characteristics.

As a baseline and principal parameter in clinical setting, we first correlated BMI to CT derived measurements SAT, VAT as well as sarcopenia (**Figure 1h-i**). While higher BMI generally reflected greater adiposity, the analysis revealed substantial variability in fat distribution and muscle mass. Notably, sarcopenia was present even among overweight and obese patients in males highlighting the limitations of BMI in capturing body composition. We then correlated overweight, reflected by BMI ≥25.0 kg/m^2^, to survival outcomes (**Figure 1j-k**): Lower BMI was associated with shorter OS (HR = 3.38, 95% CI: 1.05–11.01, p=0.032), while PFS was unaffected.

### Impact of adipose tissue distribution and Sarcopenia on outcomes after CAR T-cell infusion

We next examined prognostic markers associated with survival outcomes in our real-world cohort of patients. Based on initial findings linking BMI with OS, we investigated whether specific adipose tissue compartments might hold prognostic relevance. In univariate Cox proportional hazards analysis using adipose parameters as continuous variables, higher SAT was significantly associated with improved OS (p = 0.03, **Table 2**).

**Table 2:**
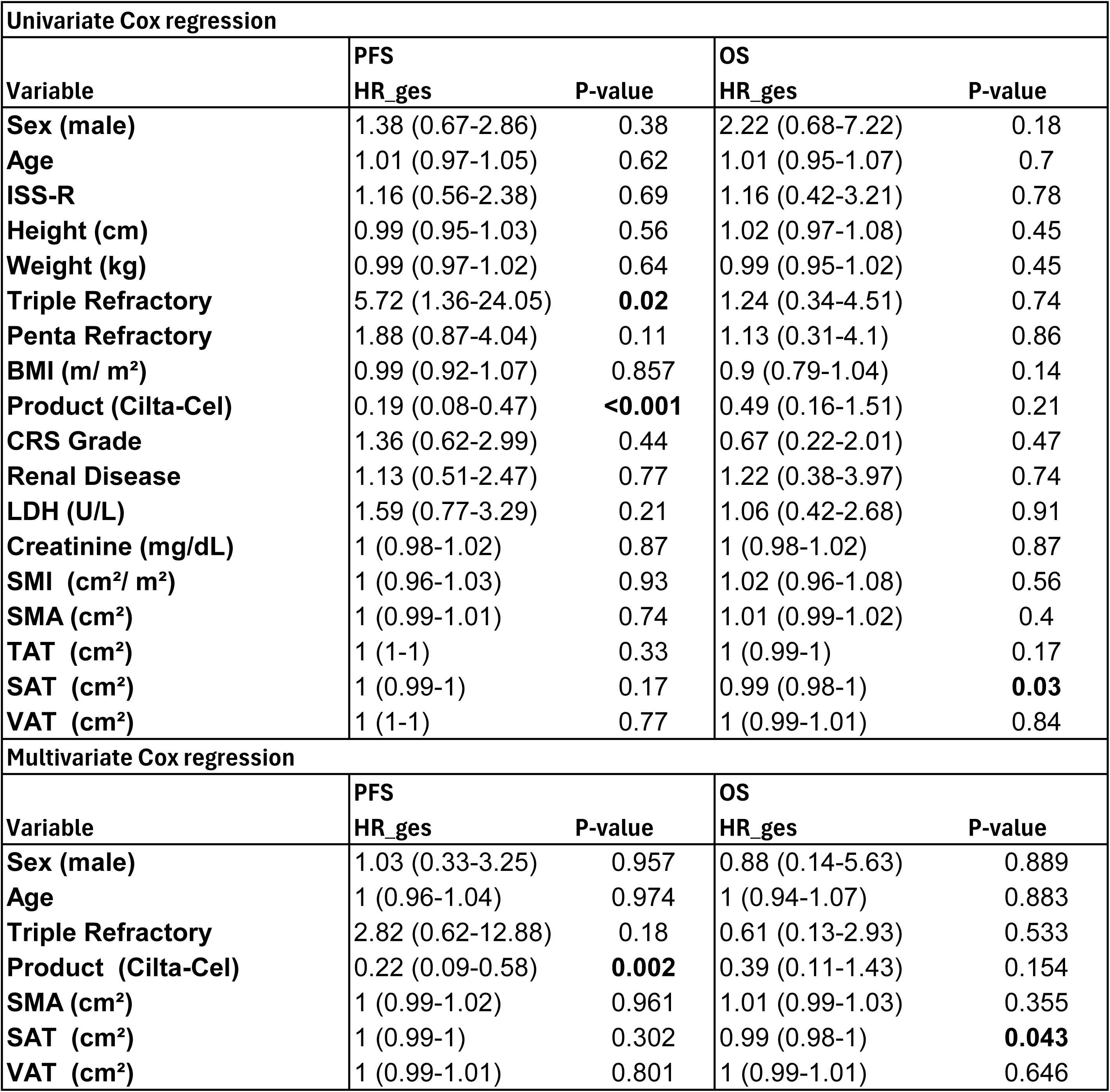

To define clinically meaningful thresholds, we used maximally selected rank statistics resulting in the following cut-offs: 227.24 cm² for TAT, 118.85 cm² for SAT, and 108.39 cm² for VAT. One patient (0.9%) was excluded from survival analysis due to insufficient follow-up. Patients with low TAT had significantly reduced OS (HR = 4.02, 95% CI:1.35–11.98, p_log-rank_ = 0.0069), while PFS was unaffected (**Figure 2a-b**). This effect that was primarily driven by SAT (HR = 4.45, 95% CI:1.45–13.63, p_log-rank_ = 0.0042; **Figure 2d**), whereas no significant differences were observed in patients with low VAT (HR = 2.54 95% CI:0.85-7.578, p_log-rank_ = 0.083; **Figure 2f**) with PFS remaining unchanged (**Figure 2c,e**).

**Fig. 2.**
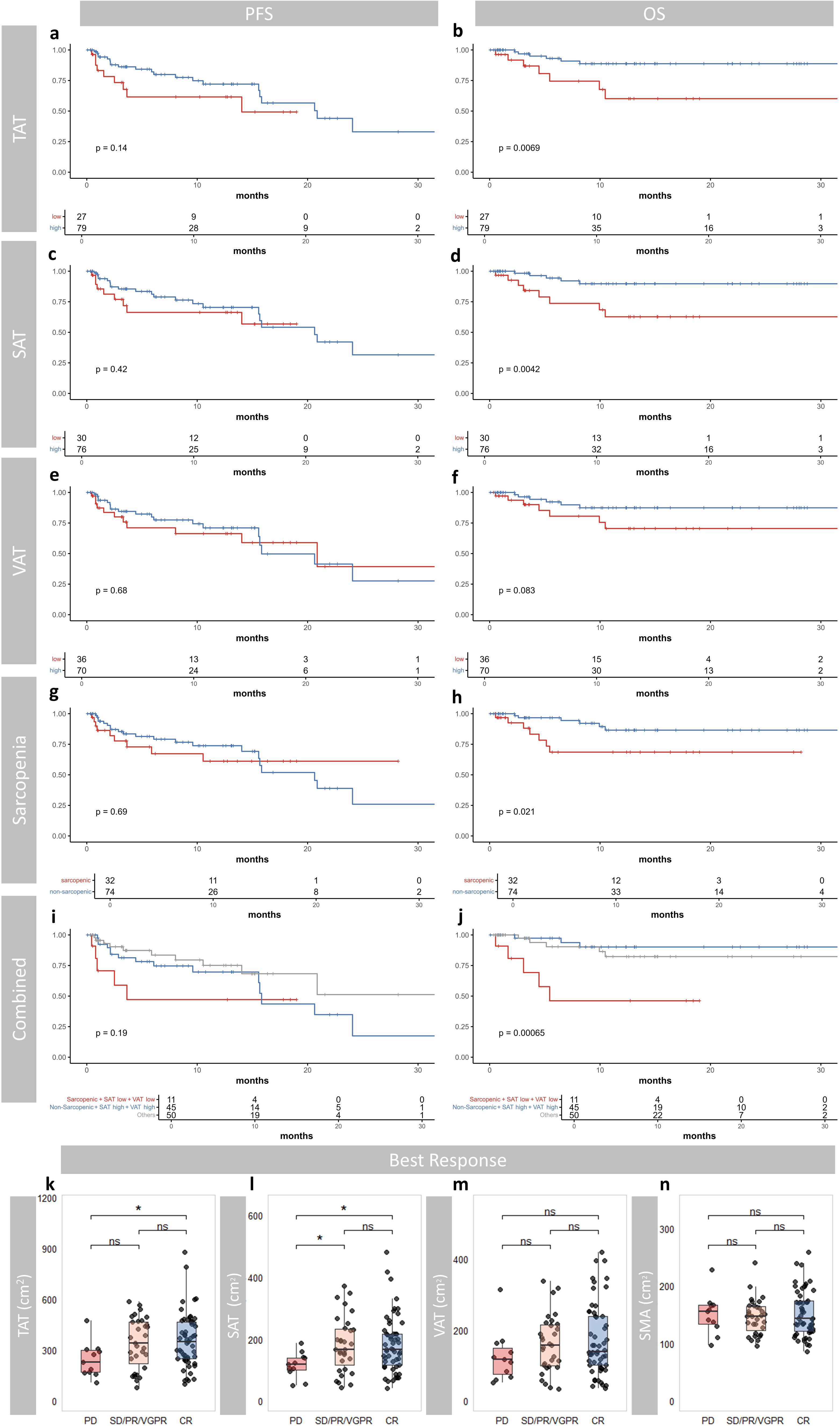
Body composition parameters and their association with survival and clinical response outcomes. **a–j**, Kaplan-Meier curves display progression-free survival (PFS, left panels) and overall survival (OS, right panels) stratified by high (blue) versus low (red) levels or non-sarcopenic (blue) versus sarcopenic (high) status of body composition parameters: (a, b) total adipose tissue (TAT), (c, d) subcutaneous adipose tissue (SAT), (e, f) visceral adipose tissue (VAT), and (g, h) sarcopenia status. (i, j) Combined stratification integrates sarcopenia status with SAT and VAT levels, comparing the high-risk group (Sarcopenic + SAT[low] + VAT[low]) against the reference category (Sarcopenic + SAT[high] + VAT[high]) and other combinations. Numbers at risk are indicated below each plot. p-values are derived from log-rank tests. **k–n**, Boxplots illustrate distributions of (k) TAT, (l) SAT, (m) VAT and (n) SM across best clinical response categories, with statistical comparisons performed using Wilcoxon rank-sum tests (significance indicated as *p < 0.05; ns = not significant).

In line with these findings, patients with lower TAT and SAT values showed inferior overall treatment responses. Adipose tissue distribution inversely correlated with depth of response, with the highest values in complete responders (CR) and the lowest in patients with progressive disease (PD) (TAT: PD vs. CR, p < 0.05; SAT: PD vs. SD/PR/VGPR and PD vs. CR, p < 0.05; **Figure 2k–m**).

In a multivariate Cox regression model adjusting for relevant confounders including SAT, VAT, SMA, age, product, refractoriness and sex, Cilta-cel treatment was independently associated with prolonged PFS (p = 0.002). Consistent with earlier findings, low SAT remained independently associated with inferior OS (p = 0.043), supporting its potential as a robust prognostic marker (**Table 2)**.

Next, we assessed the prognostic role of sarcopenia defined by established SMI thresholds (<52.4 cm²/m² for men and <38.5 cm²/m² for women) (22). While sarcopenia did not affect PFS (**Figure 2g**) or treatment response (**Figure 2n**), OS was significantly reduced in sarcopenic patients compared to non-sarcopenic counterparts (HR = 3.35, 95% CI: 1.13–9.9, p_log-rank_ = 0.021, **Figure 2h**). No significant differences were observed in uni- and multivariate analyses using SMA as a continuous variable (**Table 2)**.

We then stratified patients by sarcopenia and adipose tissue parameters grouping those with both low SAT and low VAT (sarcopenic+SAT[low]+VAT[low]) against non-sarcopenic individuals with high SAT and high VAT (sarcopenic+SAT[high]+VAT[high]), as well as all remaining combinations (**Figure 2i-j**). The sarcopenic+SAT[low]+VAT[low] cohort exhibited markedly poorer OS compared to the sarcopenic+SAT[high]+VAT[high] group (HR 8.83, 95% CI 2.18– 37.26.6, p = 0.0028) and compared to the remaining groups combined (HR 5.01, 95% CI 1.47–17.63, p = 0.0101), whereas no differences were noted for PFS.

Collectively, these results establish SAT and sarcopenia as prognostic markers for OS following CAR T-cell therapy.

### Sarcopenia and low SAT are associated with reduced bystander T-cell levels at leukapheresis

To investigate mechanisms behind the differences in OS and response, we evaluated the impact of TAT, SAT, VAT, and sarcopenia on T-cell differentiation and CAR T-cell expansion from LP to day 100 post-infusion. Patient and sample counts are shown in **Figure 3a**.

**Fig. 3.**
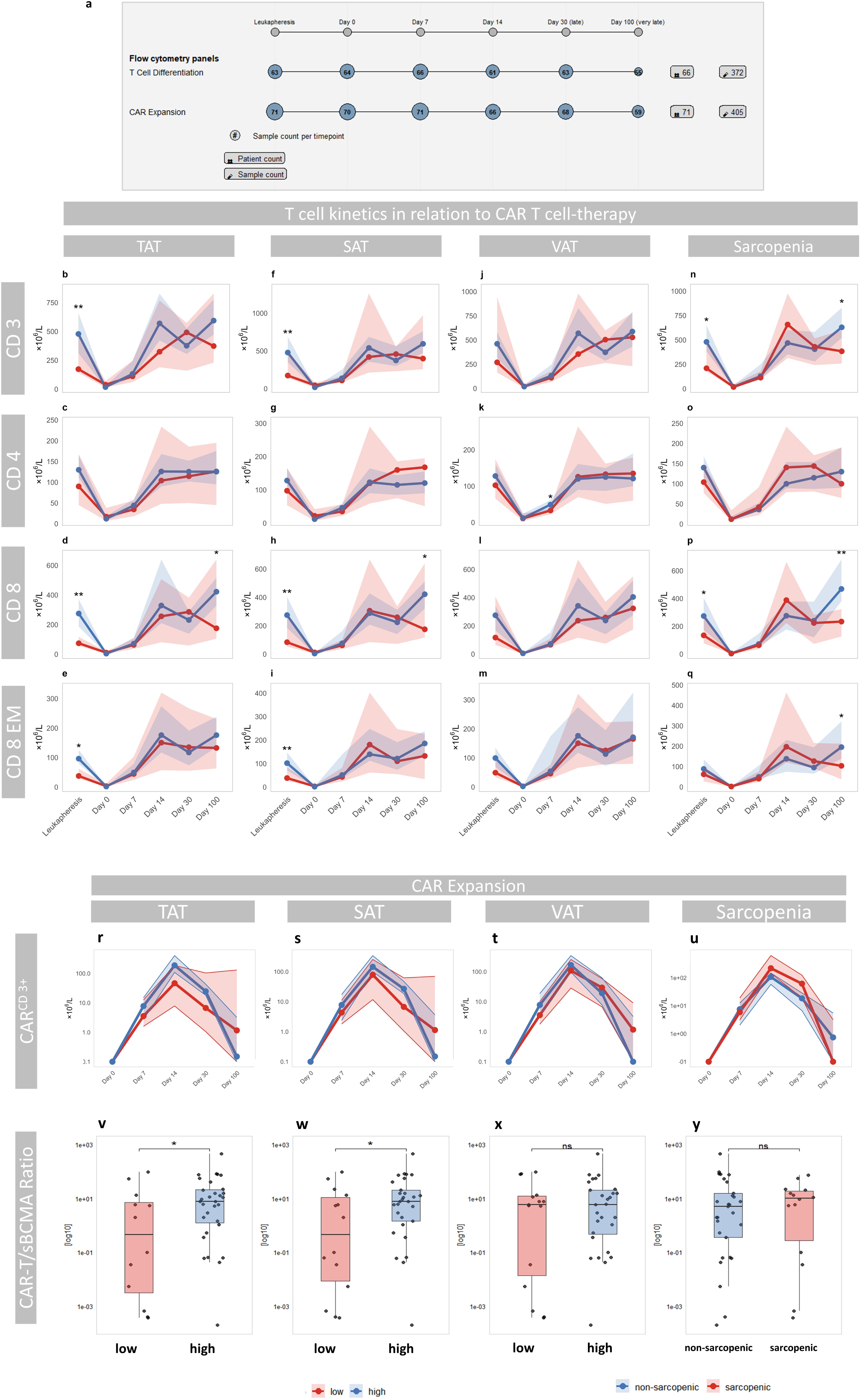
T-cell kinetics and Effector-Target ratio in relation to body composition parameters. **a**, Schematic overview of the study design, highlighting the time points of leukapheresis and post-infusion sampling (LP, Day 0, 7, 14, 30, and 100), along with the number of patients and samples included in T-cell differentiation and CAR T-cell expansion panels at each time point. **b–q**, display total CD3⁺, CD4⁺, CD8⁺ and CD8⁺ effector memory (EM) T-cell counts over time, stratified by TAT (b-e), SAT (f-i), VAT (j-m) and sarcopenic status (n-q). **r-u**, total CD3⁺ CAR T-cell Expansion across CAR T-cell therapy stratified by by TAT (r), SAT (s), VAT (t) and sarcopenic status (u). **v-y**, CD3⁺ CAR T-cell/ sBCMA-Ratio calculated at day 30 post infusion stratified by TAT (v), SAT (w), VAT (x) and sarcopenia status (y). In all plots, red and blue lines represent patients with low vs. high adiposity, or sarcopenic vs. non-sarcopenic status, respectively; shaded areas indicate the 95% confidence intervals with statistical comparisons performed using exact one-sided Wilcoxon rank-sum tests. (significance indicated as *p < 0.05; **p < 0.01, ns = not significant). For longitudinal group comparisons, *p*-values were adjusted for multiple testing at each time point using the Benjamini–Hochberg method.

At LP, patients with low TAT and SAT exhibited reduced counts of CD3⁺ (p < 0.01), CD8⁺ (p < 0.01), and CD8⁺ effector memory (CD8⁺EM) T cells (p < 0.05, **Figure 3b–i**). While no differences were observed during early follow-up CD8⁺ levels were increased at day 100 after infusion (p<0.05). In contrast, VAT was not associated with differences in T-cell dynamics, except for reduced CD4⁺ levels at day 7 (p<0.05, **Figure 3j–m**). Similarly to SAT, sarcopenic patients had lower CD3⁺ (p<0.05) and CD8⁺ (p<0.05) counts at LP as well as reduced CD3⁺ (p<0.05) and CD8⁺ (p<0.01) levels at day 100 (**Figure 3n–q**).

CAR T-cell expansion was not affected by adipose tissue parameters or sarcopenia, with no significant differences observed across TAT, SAT, VAT, or in sarcopenic patients (**Figure 3r–u**). However, at day 30 post-infusion, patients with low TAT (p = 0.01) and SAT (p = 0.05) exhibited both elevated absolute sBCMA levels (**Table 1**) as well as a significantly decreased effector-to-target ratio-defined as CAR T-cell counts relative to sBCMA levels (TAT: p<0.05, SAT: p<0.05, **Figure 3v-y**).

These findings suggest that low SAT and sarcopenia at baseline are associated with significant differences in T-cell dynamics and differentiation as well as a diminished reduction of tumor burden following CAR T-cell therapy. At least for low SAT this effect can be traced back to an impaired effector-to-target ratio.

### Sarcopenia and Adipose Depletion Drive T-Cell Transcriptional Remodeling

Since flow cytometry showed significant differences in the T-cell compartment based on body composition, we next investigated its impact on quantitative and qualitative cellular dynamics following CAR T-cell infusion at a single-cell resolution. We included 75 patients from our study cohort with available single-cell data from peripheral blood to gain mechanistic insights into how body composition affects MM physiology and CAR T-cell function (**Figure 4**). In total, approximately 600,000 cells from 178 samples (median: 3050 cells per sample, range: 554–8471; **Figure 4a**) from LP, Late and Very late timepoints were included in the analysis.

**Fig. 4.**
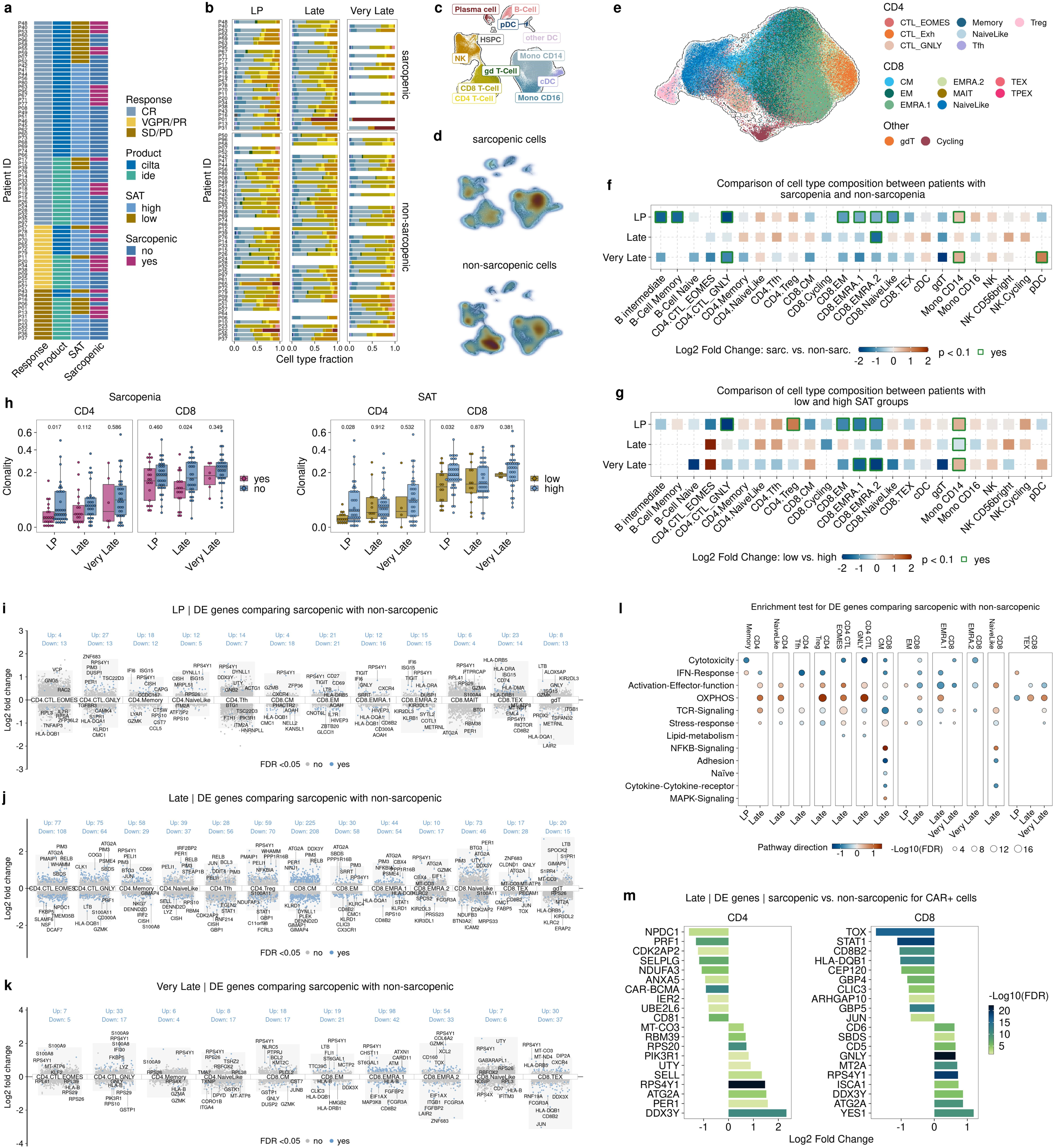
Longitudinal profiling of BCMA-targeting CAR T-cell therapy in multiple myeloma at single-cell resolution. **a**, Overview of sample availability for single-cell analysis. **b**, Bar graphs show summary statistics of the composition of cell types for each sample time point. Around 600,000 cells from 178 samples were analyzed. Cell type color codes are consistent with **c**, which depicts the Uniform Manifold Approximation and Projection (UMAP) of the cells. **d**, UMAP view of cell densities in individuals with and without sarcopenia. High relative cell density is shown as dark red. **e**, Approx. 200,000 T-cells were embedded using the UMAP method. Cells are colored according to T-cell subtypes. **f-g**, Differences in cell type composition between patients with and without sarcopenia (f) and between SAT low and high (g). For each time point and cell type, the log2 fold change (log2FC) in mean cell fraction between sarcopenic and non-sarcopenic samples (f) and between SAT low and high (g) was calculated and color-coded. A positive log fold change indicates a higher cell type fraction in patients with sarcopenia or SAT low. Significant differences (unadjusted p-values) were estimated using empirical Bayes moderated t-statistics (two sided) implemented in the speckle package (*p = 0.1, **p = 0.05, ****p = 0.001, *****p < 0.0001). **h**, Based on TCR-Seq, differences in CD4⁺ and CD8⁺ T-cell clonality between individuals with and without sarcopenia are depicted. Clonality was assessed by grouping cells according to their clonotype size, allowing comparison of repertoire diversity across conditions. **i-k**, Differential gene expression analysis for T-cell subtypes comparing sarcopenic with non-sarcopenic samples. Only subtypes with significant (adjusted p-value < 0.05) changes in gene expression are shown. Shown are the highest ranked (sorted by log2FC) protein-coding DE genes. **l**, Enrichment analysis for DE genes from all time points for T-cell subtypes. The dot plot depicts enriched T-cell signatures (adjusted p-value < 0.05). The color indicates the pathway direction, which is the number of DE genes with a log fold change of >0 minus the number of DE genes with a log fold change of <0 divided by the square root of the number of pathway-associated genes. **m**, Differential gene expression analysis for CD4 and CD8 CAR^+^ cells comparing sarcopenic with non-sarcopenic samples for the Late time point. Shown are the highest ranked (sorted by log2FC) protein-coding DE genes with a log2FC > 0 and < 0. A positive log fold change indicates upregulation in patients with sarcopenia.

Based on the above-mentioned findings, which identified SAT and sarcopenia as the most relevant parameters, we focused exclusively on these two variables for downstream single-cell analyses. An overview of the sample availability is shown in **Figure 4a**, while cell type composition and annotations on coarse level are shown in **Figure 4b–d** for each sample. All analyzed T-cell subtypes are depicted in **Figure 4e**. For SAT, 65 samples were available at LP (16 low and 49 high), 66 at Late (17 low and 49 high), and 47 at Very Late time point (8 low and 39 high). Regarding sarcopenic status, 65 samples were included at LP (41 non-sarcopenic and 24 sarcopenic), 66 at Late (41 non-sarcopenic and 25 sarcopenic), and 47 at Very late time point (36 non-sarcopenic and 11 sarcopenic).

Analysis of cell type abundancies revealed that sarcopenic patients demonstrated a significant reduction in B- and T-cell subsets, including B intermediate cells, memory B-cells, granulysin-expressing cytotoxic CD4^+^ (CD4^+^CTL_GNLY), CD8^+^EM, CD8^+^ EM re-expressing *CD45RA* 1 (CD8^+^EMRA.1), CD8^+^EMRA.2, and CD8^+^naive-like cells (**Figure 4f, Figure S1**). These differences were most pronounced at LP. In contrast, CD14⁺ monocytes were significantly increased in sarcopenic compared to non-sarcopenic patients at LP. At later time points, sarcopenic individuals showed persistently altered immune profiles, with reduced CD4.CTL_GNLY cells and increased CD8^+^EMRA.2 as well as plasmacytoid dendritic cells (pDCs, **Figure 4f**).

Findings from SAT depleted patients were consistent with results from sarcopenic patients: We observed lower frequencies of CD4^+^CTL_GNLY, CD8^+^EM, CD8^+^ EMRA.1, and CD8^+^EMRA.2 cells, while CD4⁺ regulatory T-cells (Tregs) and monocyte CD14⁺ cells were increased compared to patients with high SAT at LP (**Figure 4g, Figure S2**). Differences in CD8^+^EMRA.1 and CD8^+^EMRA.2 cell populations were also present at Very late time point.

After identifying significant differences in the T-cell compartment at LP and the first 100 days following CAR T-cell infusion, we analyzed TCR clonality. In general, CAR T-cell therapy was associated with a progressive increase in TCR repertoire clonality from LP to Late and continuing through Very Late time point post-infusion. Further assessment of the TCR repertoire showed that sarcopenic patients exhibited reduced CD4⁺ T-cell clonality at LP and decreased CD8⁺ T-cell clonality at the Late time point (**Figure 4h, Figure S3**). In patients with low SAT, clonality of both CD4⁺ and CD8⁺ T-cells was consistently reduced at LP, with no notable changes during follow-up.

Building on these compositional and clonal insights, we next sought to define the transcriptional programs underlying T-cell dysfunction in sarcopenia and low SAT.

**Figure 4i-k** shows the significantly (FDR <0.05) differentially expressed (DE) genes comparing sarcopenic with non-sarcopenic patients for each subtype and time point (**Figure S4**). Notably, at the late time point, CD8^+^ central memory (CM) cells in sarcopenic patients showed the greatest transcriptional perturbation, with 225 genes upregulated and 208 downregulated DE genes. Enrichment analysis revealed broad downregulation (FDR < 0.05) of key immune pathways, including cytotoxicity, interferon (IFN) and stress response, across both CD4⁺ and CD8⁺ lineages. In CD8⁺ cells, activation and effector functions were largely suppressed, whereas CD4⁺ cells showed upregulation of these pathways. TCR signaling exhibited a mixed pattern (**Figure 4l**). In sarcopenic patients, oxidative phosphorylation (OXPHOS) was significantly enriched in CD4^+^CTL_GNLY, eomesodermin (EOMES) expressing CD4^+^CTL (CD4^+^CTL_EOMES), CD4^+^ naive like, CD4^+^ memory cells and CD4⁺ Treg cells as well as CD8^+^EM, CD8^+^ EMRA.1 and exhausted CD8^+^ cells (CD8^+^TEX) after CAR T-cell infusion, indicative for mitochondrial dysfunction (26,27), whereas NF-κB signaling was enhanced in CD8^+^CM and CD8^+^ naive like cells.

Stratification by SAT mirrored these patterns (**Figure S5**). In patients with low SAT all subsets exhibited downregulation of cytotoxicity and activation–effector function after treatment, except for CD4⁺ naive-like and CD8 naïve-like cells, which showed increased activation–effector function. OXPHOS pathway was upregulated in CD4^+^CTL_GNLY, CD4^+^CTL_EOMES, CD8^+^CM, CD8^+^EM, CD8^+^EMRA.1 and CD8^+^TEX cells after infusion and NF-κB activity was increased in CD8^+^ naive-like cells.

The significant alterations in the T-cell compartment at LP and throughout the first 100 days post-infusion led us to hypothesize that both variability in the input T-cell material for CAR T-cell manufacturing and subsequent T-cell perturbations may confer functional differences in CAR T-cell qualitative properties, modulated by sarcopenia. DE analysis at the late time point, comparing CAR⁺ T-cells from sarcopenic versus non-sarcopenic patients, revealed downregulation of cytotoxic (PRF1), cell cycle (CDK2AP2), and adhesion (SELPLG) genes as well as the BCMA-CAR construct in CD4⁺ cells. Decreased expression of exhaustion markers (TOX, STAT1) and checkpoint genes (HLADQB1, CD8B2) was observed in CD8⁺ cells. At the same time upregulation of mitochondrial (MTCO3) and ribosomal (RPS20) genes in CD4^+^ cells, autophagy gene (ATG2A) in CD4^+^ and CD8^+^ cells, and cytotoxic effector genes (GNLY, CD5, CD6) in CD8^+^ cells (**Figure 4m**).

Taken together, these data reveal that both sarcopenia and low SAT impose a multifaceted (CAR) T-cell dysregulation, characterized by reduced cell counts at LP, concomitant suppression of cytotoxic responses, and enhanced activation of inflammatory and mitochondrial pathways following CAR T-cell infusion.

## Discussion

In this study, we systematically assessed the impact of currently used body composition parameters in the context of anti-BCMA directed CAR T-cell therapy for RRMM. In our cohort, we observed that low levels of TAT and SAT are associated with reduced OS and diminished therapeutic response as well as reduced OS for sarcopenic patients. Furthermore, low SAT and sarcopenia were linked to reduced bystander T-cell repertoire at LP. Single-cell analysis confirmed reduced immunologic activity by diminished T-cell compartments and revealed lower clonality at LP and shared immunometabolic reprogramming.

Generally, fat distribution and sarcopenia represent distinct but equally adverse metabolic states in cancer, both associated with poor prognosis and impaired therapeutic success (9,16,23,28). Here, CT-derived body composition metrics are increasingly used to refine risk stratification across tumor types (9,13,24,29–31). VAT promotes inflammation and cancer progression (30,32) and correlates with poor survival in solid tumors, though its role in hematology is controversially discussed (7,13,33). SAT demonstrates a context-dependent prognostic role that varies across cancer types. While high SAT levels have been associated with poor outcomes in solid tumors such as non-small cell lung cancer (31), they have also emerged as prognostic factors in hematologic malignancies. For instance, in lymphoma, lower levels of TAT, SAT, and VAT were consistently linked to inferior PFS and OS (7).

Regarding MM, we found that BMI < 25 kg/m² and low TAT, primarily driven by SAT, were associated with inferior OS, diminished response rates and elevated sBCMA levels pre- and post-treatment, indicative for increased tumor burden (34). While the protective prognostic value of increased BMI in MM has been demonstrated in a study of nearly 3,000 newly diagnosed patients (35), the value of SAT remains inconclusive, and current knowledge is restricted to newly diagnosed disease. One study reported no association between SAT or VAT and survival after autologous stem cell transplantation (ASCT) at 2 years (36), while others suggest VAT may predict outcomes over 10 years (13). Our findings align with reports associating low SAT with worse survival, whereas results for VAT showed no impact (15). PET/CT studies indicate high SAT radiodensity signals poor prognosis, possibly due to increased metabolic activity and inflammation (33). Notably, higher radiodensity correlated with lower SAT and VAT volumes, indicating a hypermetabolic, aggressive disease phenotype. This is further supported by our findings that low TAT and SAT were associated with reduced response rates and increased tumor burden as shown by sBCMA assessment.

Differences between the findings from ASCT studies and our current results may be explained by the earlier application of ASCT in newly diagnosed multiple myeloma (NDMM) compared with the use of CAR T-cell therapy in heavily pre-treated RRMM, as well as by their distinct toxicity profiles (37,38).

Patients with low SAT showed fewer bystander T-cells at LP. Single-cell analysis confirmed reduced key effector subsets, matching flow cytometry data. An immunosuppressive microenvironment characterized by low CD8⁺ T-cell counts and reduced functionality at LP has previously been associated with poor outcomes (39), while reduced T-cell clonality in both CD4⁺ and CD8⁺ compartments correlate with lower response rates after CAR T-cell therapy (5) or treatment with bispecific antibodies (40). These findings are consistent with worse responses observed in low SAT patients in our current study. Metabolically, low SAT patients exhibited upregulation of mitochondrial OXPHOS pathway after infusion, rendering T-cell exhaustion and impaired function (26,27). Notably, NF-κB pathway activation was visible in CD8 naïve like cells suggesting inflammatory conditions (41).

These findings suggest that low SAT not only reflects reduced metabolic reserves but also impaired immune competence and diminished CAR T-cell efficacy. Among adipose compartments, SAT emerged as the most consistent predictor of OS, whereas VAT, despite showing trends toward survival, demonstrated no clear associations with other parameters. This contrasts with observations in lymphoma and solid tumors, where both SAT and VAT have been reported to hold broader prognostic significance (7). These differences may reflect methodological variation or disease-specific biology, emphasizing the context-dependent nature of adipose tissue in cancer prognosis-often described as the obesity paradox (28). Our results underscore the importance of interpreting body composition within specific disease contexts and suggest that, although increased VAT generally carries negative implications even outside oncologic settings, low VAT in this context may instead reflect poor nutritional status. This assumption is supported by our analysis showing that sarcopenic patients, who also exhibited low SAT and VAT values, experienced the poorest outcomes.

Sarcopenia is a well-established prognostic factor in various solid tumors (9,42,43) and has shown adverse implications in AML (44,45) and ALL (46). In ASCT, sarcopenia showed no significant impact on survival, though trends were observed over 10 years (13). In the specific setting of anti-CD19 CAR T-cell therapy, sarcopenia has been identified as a reliable predictor of worse OS, but not PFS in lymphoma patients (7), consistent with our findings. This negative effect is thought to stem from a diminished physiological reserve, impairing the patient’s ability to tolerate toxic therapies such as LDC (47) or subsequent lines of salvage therapies. In our data, this is reflected in impaired immune function at LP, including numerical reductions in B- and T-cell subsets, increased monocyte fraction, reduced CD4⁺ clonality, and downregulation of key immune pathways responsible for cytotoxicity, IFN and stress response with equal possible implications adverse events as for SAT (5,39,40,48). The concept of reduced functional reserve in sarcopenic patients has also been described in immune checkpoint inhibitors (49). Strikingly and in line with our observations in low SAT patients, sarcopenic patients underwent upregulation of OXPHOS pathway after CAR T-cell infusion.

Beyond their prognostic value, adipose tissue parameters-especially SAT- and sarcopenia are potentially modifiable factors with important therapeutic implications. Poor nutritional status and cancer-associated cachexia at baseline-defined as loss of skeletal muscle and adipose tissue-have been identified as adverse prognostic indicators for survival after CAR T-cell therapy (50), possibly due to a distinct macro- and microenvironment causing systemic inflammatory processes (51). Moreover, also obesity may influence disease progression, including the transition from MGUS to MM (16). Interventions such as structured physical activity facilitate the T-cell/muscle interaction improving immune functionality (52). In addition, exercise, nutritional surveillance, and the judicious use of broad-spectrum antibiotics not only preserve muscle mass and metabolic health but also favorably reshape the gut microbiome (53), which represents an emerging regulator of systemic immunity and CAR T-cell expansion (54). Fecal microbiome sequencing data have linked specific microbial profiles, including *Faecalibacterium prausnitzii, Bifidobacterium*, and other butyrate producers, to improved CAR T-cell functionality and reduced toxicity (3,54). In turn, microbial dysbiosis is common in sarcopenia (55), and malnutrition as well as lack of physical activity may impair CAR T-cell efficacy. Thus, modulating the microbiome using these different approaches could be a promising adjunct to optimize CAR T-cell therapy outcomes.

This study has several limitations. First, it is a retrospective, single-center analysis with a relatively short median follow-up, limiting comparability to long-term outcome studies (13). Second, no follow-up imaging was available after CAR T-cell therapy, preventing longitudinal assessment of body composition changes as performed by other groups (47). Additionally, there is no consensus on cut-off values for adipose tissue compartments, and sarcopenia definitions are derived from solid cancer potentially limiting generalizability (23).

In summary, SAT and sarcopenia are important prognostic factors associated with OS in RRMM patients undergoing anti-BCMA CAR T-cell therapy. Their association with altered immune cell composition at LP, diminished CAR T-cell/sBCMA ratio and metabolically disrupted immune pathways suggests a mechanistic link to reduced therapeutic efficacy. Notably, these parameters can be reliably quantified from routine cross-sectional imaging, enabling seamless integration into longitudinal clinical monitoring without added diagnostic burden. Given their modifiable nature, interventions such as personalized nutrition and exercise, these therapies hold promise to improve outcomes. Additionally, moving beyond BMI as a sole measure, as recently recommended by an international expert commission, may improve patient stratification and therapeutic decision-making (56). Prospective multicenter studies are needed to validate these findings and define standardized thresholds for clinical implementation.

## Acknowledgements

This work was funded by grants from the International Myeloma Society (IMS Research Grant 2023) and the German Research Foundation (SPP µbone) and EU HORIZON (CERTAINTY). The CERTAINTY project is funded by the European Union (Grant Agreement 101136379). Views and opinions expressed are however those of the authors only and do not necessarily reflect those of the European Union. Neither the European Union nor the granting authority can be held responsible for them.

The authors acknowledge the financial support by the Federal Ministry of Education and Research of Germany and by Sächsisches Staatsministerium für Wissenschaft, Kultur und Tourismus in the programme Center of Excellence for AI-research “Center for Scalable Data Analytics and Artificial Intelligence Dresden/Leipzig”, project identification number: ScaDS.AI

Figure 1a “Study Workflow” was created in BioRender. Rade, M. (2026) https://BioRender.com/zcp7oln

Authors express their gratitude towards patients and their families who participated in our research.

## Author contributions

TCW analyzed data, designed figures and took lead in writing the manuscript.

MR analyzed data and designed figures.

NG and MF analyzed data.

HJM and TiD assessed body composition parameters radiologically.

DF, PB, LF, SS, AG, MaF, RB, MK, KHM, MH, CDH, MJ, GNF, AB managed patient care and data collection.

TN, UAS, TD, UK, KR, UP, VV, HJM and MM supervised the project.

HJM and MM devised the main conceptual ideas and revised the manuscript.

All authors discussed the results have approved of the manuscript.

## Data availability

The data used in this study is available upon reasonable request to

---

**Fig. S1 | Comparison of cell types for sarcopenic and non-sarcopenic individuals for each time point.**

**a-c,** Differences between cell type proportions were evaluated using the speckle R package. Significant differences (unadjusted p-values) were estimated using empirical Bayes moderated t-statistics (two sided) implemented in the speckle package (*p = 0.1, **p = 0.05, ****p = 0.001, *****p < 0.0001).

**Fig. S2 | Comparison of cell types for SAT low and SAT high groups for each time point.**

**a-c,** Differences between cell type proportions were evaluated using the speckle R package. Significant differences (unadjusted p-values) were estimated using empirical Bayes moderated *t*-statistics (two sided) implemented in the speckle package (\**p* = 0.1, \*\**p* = 0.05, \*\*\*\**p* = 0.001, \*\*\*\*\**p* < 0.0001).

**Fig. S3 | Differences in T-cell subtype clonality.**

**a,** Comparison between individuals with and without sarcopenia are depicted. Difference in between SAT high and low groups are shown in **b,** Two-sided Wilcoxon rank sum tests were performed to calculate p-values between the groups at each time point, respectively

**Fig. S4 | DGEA comparing cells from sarcopenic with non-sarcopenic individuals.**

**a,** Bar plots show custom gene modules with DE genes (adjusted p-value < 0.05) that functionally characterize T-cells.

**Fig. S5 | Comparison between patients with SAT low and SAT high.**

**a-c,** Differential gene expression analysis for T-cell subtypes and time points comparing SAT low with SAT high patients. Shown are the highest ranked (sorted by log2FC) protein-coding DE genes. A positive log2 fold change indicates upregulation in SAT low patients. **d,** Enrichment analysis for DE genes from all time points for T-cell subtypes. The dot plot depicts enriched T cell signatures (adjusted p-value < 0.05). The color indicates the pathway direction, which is the number of DE genes with a log fold change of >0 minus the number of DE genes with a log fold change of <0 divided by the square root of the number of pathway-associated genes.

