## Supplementary figures and images for "Body composition predicts poor outcomes and reveals immunometabolic dysfunction via single-cell profiling in anti-BCMA CAR T-treated myeloma"

### Supplementary Figures S1-S5

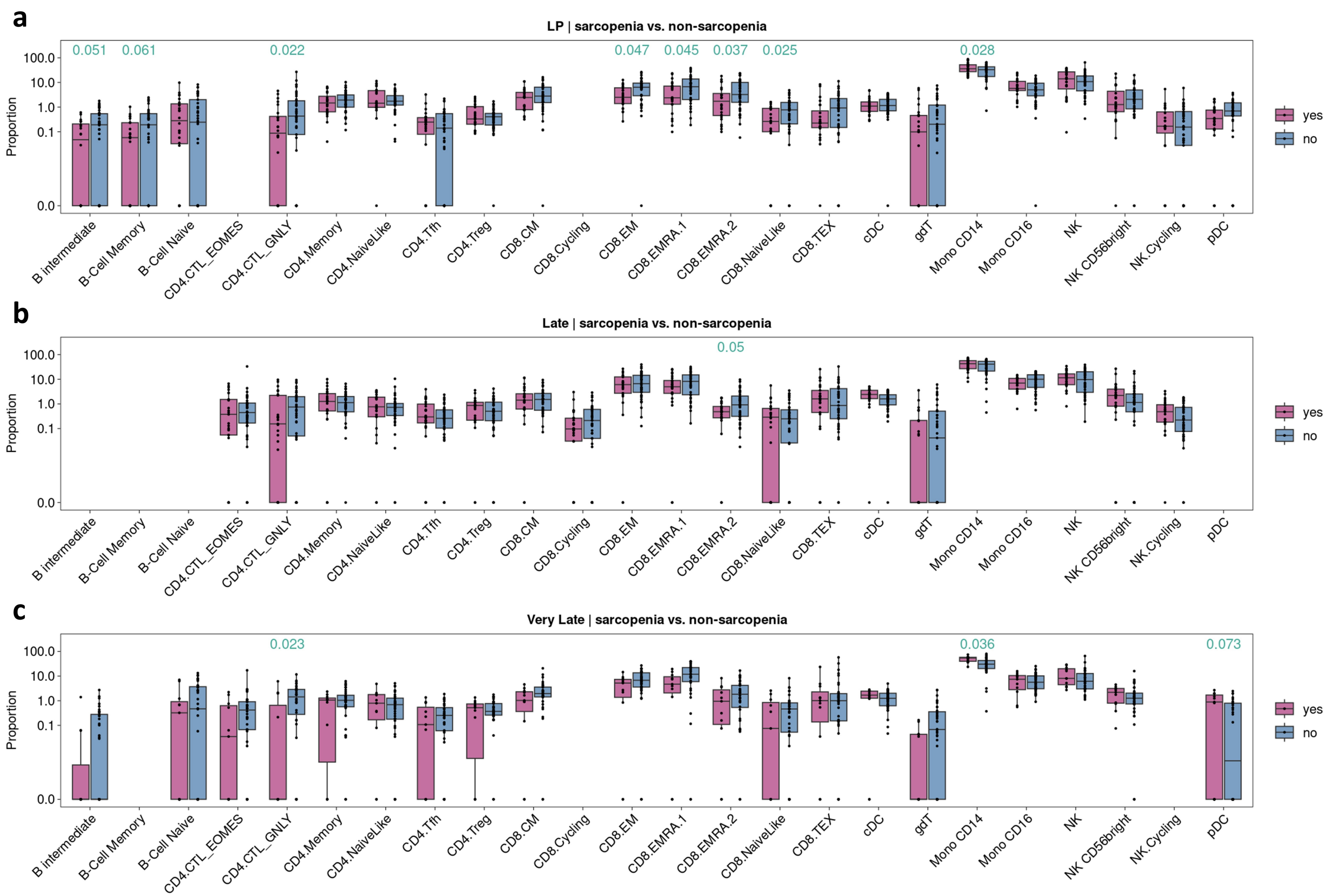

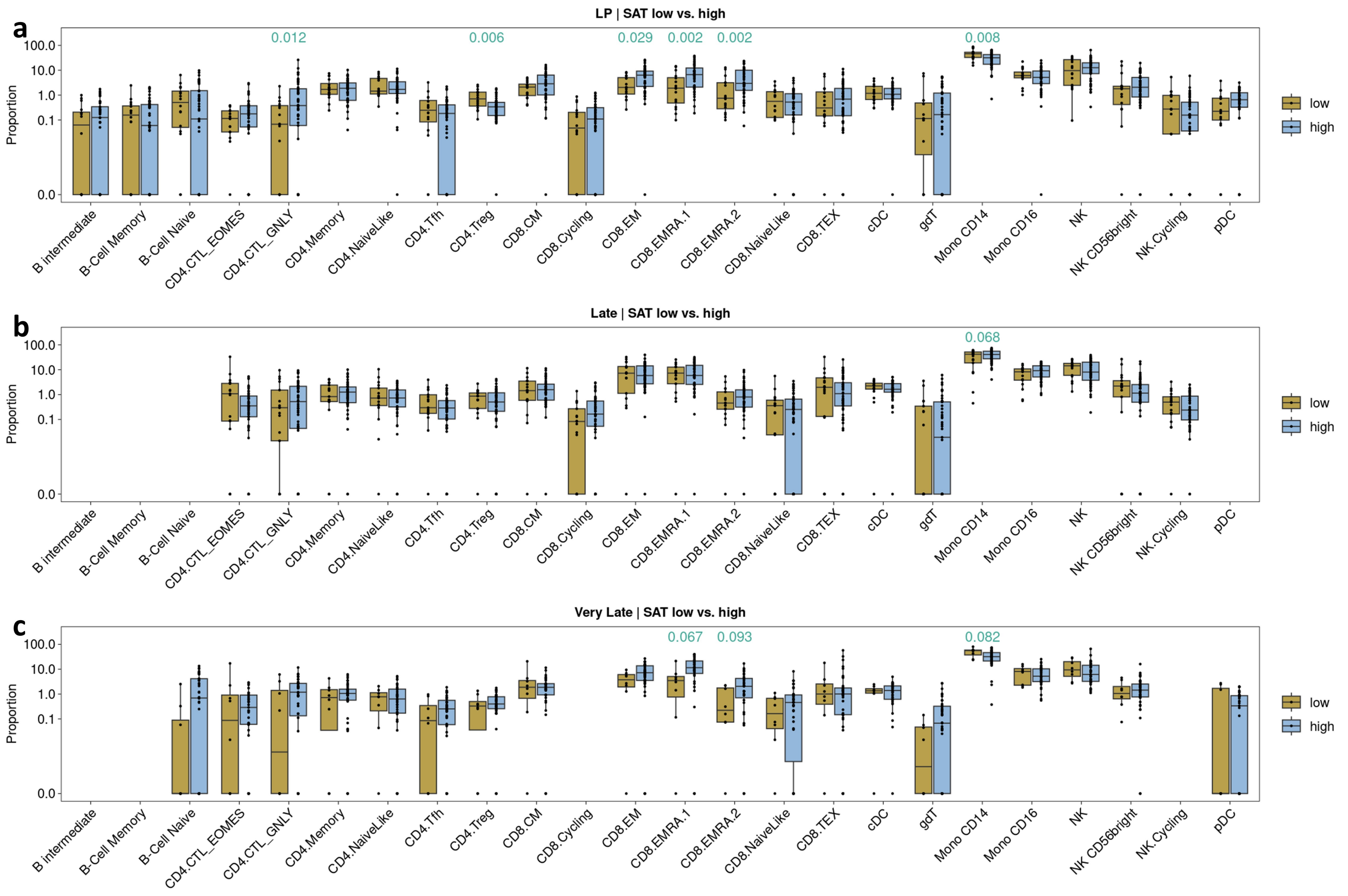

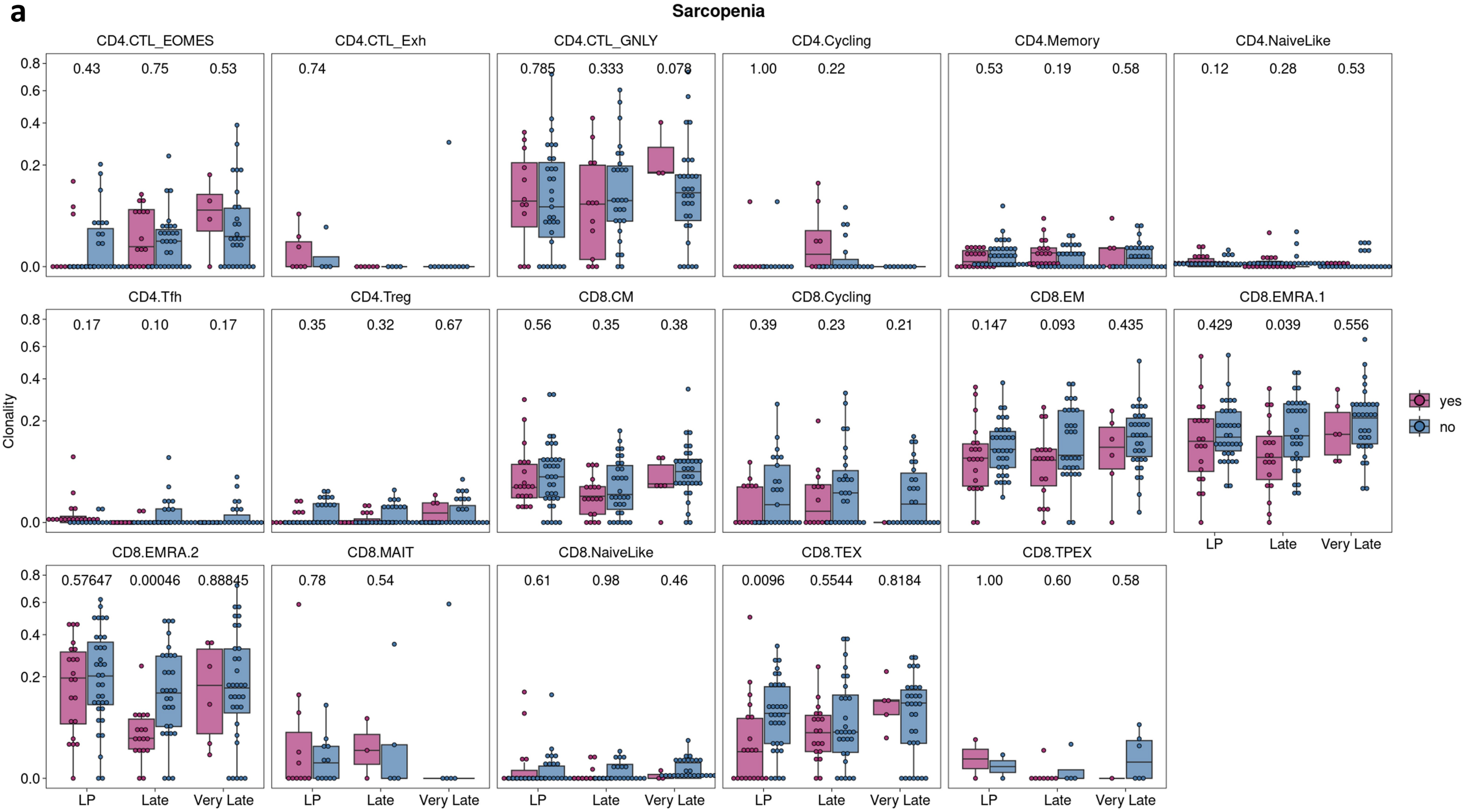

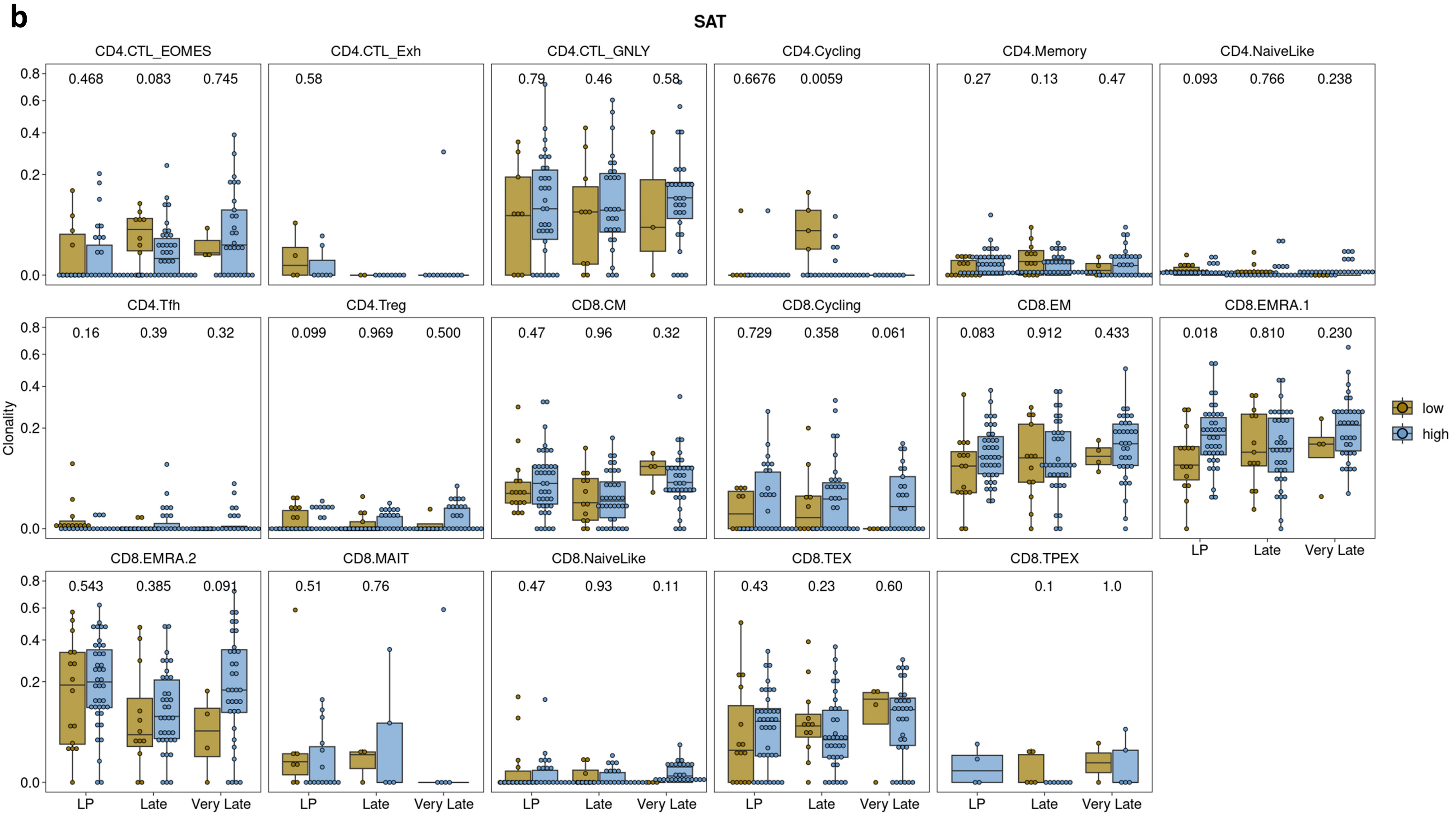

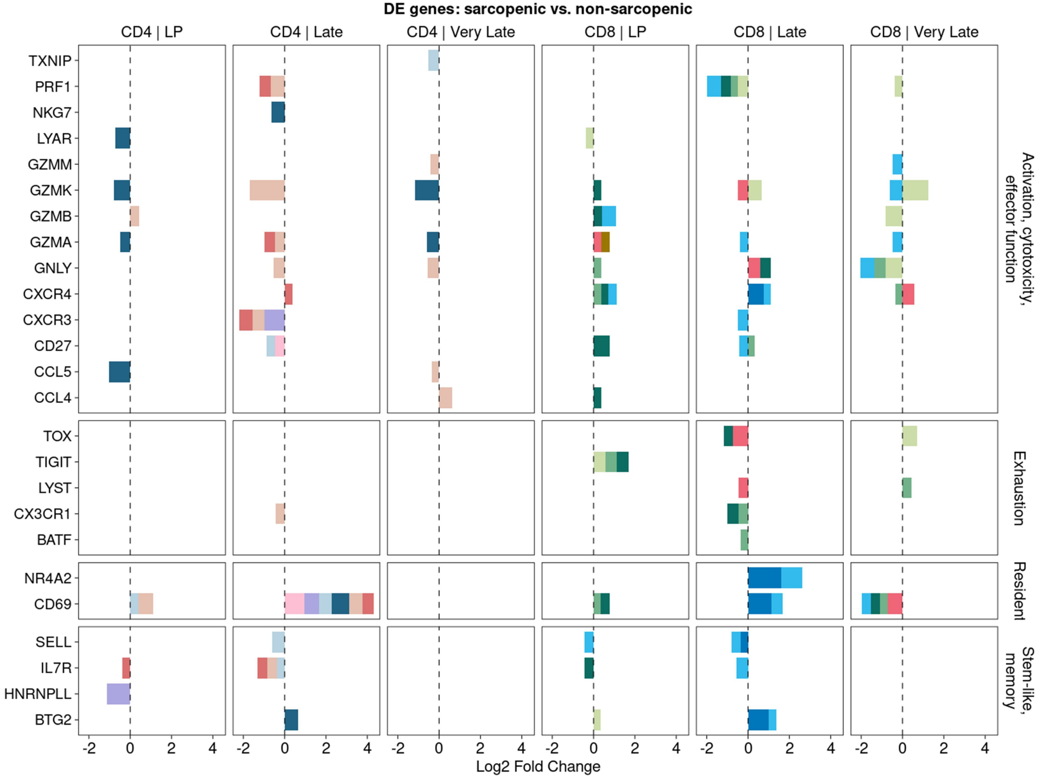

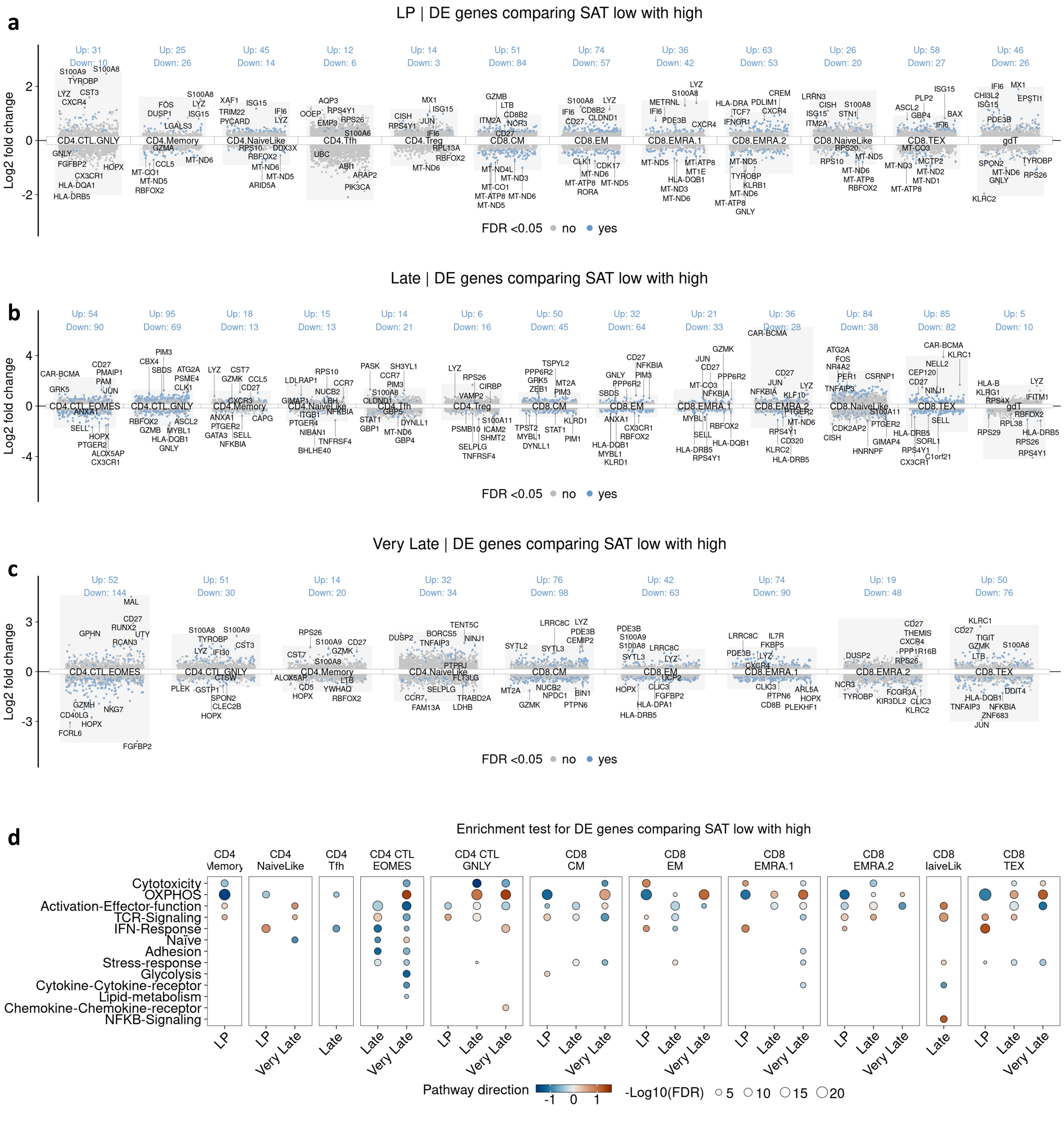
